# The prevalence of psychological distress and relationships with social fragmentation and isolation: a geospatial study and survey in rural Australia

**DOI:** 10.1101/2020.11.12.20230771

**Authors:** Victor Forcadela, Nasser Bagheri, Claudia Slimings

## Abstract

**Purpose:** There is limited data on the effects of social isolation on rural mental health. The aim of this study was to describe the prevalence of psychological distress in a rural area of Australia while exploring the association between psychological distress and social isolation at the individual and area level.

**Methods:** An online cross-sectional survey of 408 adult volunteers was conducted across rural south-eastern New South Wales from October 2019 to April 2020. The Kessler 10 was used to measure psychological distress, while area-level social fragmentation was assessed using the family (ANSFI_fam_) and mobility (ANSFI_mob_) components of the Australian Neighbourhood Social Fragmentation Index. The number of different occupations of people known socially was used to assess individual-level social isolation. Spatial analysis was performed to identify any spatial autocorrelation.

**Results:** The prevalence of high psychological distress in the sample was 29%. Using logistic regression models, there was little evidence of a relationship between high psychological distress and ANSFI_fam_ (odds ratio (OR)=0.98, 95% confidence interval (CI)=0.93–1.03), or ANSFI_mob_ (OR=1.04, 95%CI=0.99–1.09). High psychological distress also did not appear to be associated with number of occupations known socially (OR=1.00, 95%CI=0.99–1.00). There was no significant spatial autocorrelation of psychological distress or social fragmentation at the postal area level.

**Conclusion:** The results suggest that social fragmentation or isolation may not have a significant effect on psychological distress in a rural setting. Nevertheless, further investigation of the effects of social isolation on mental health in rural areas is warranted.

## Introduction

Mental health is a prominent issue that affects a significant proportion of Australians. According to the National Health Survey, one in five Australians had a mental or behavioural condition in 2017-2018 [1]. Furthermore, the 2007 National Survey of Mental Health and Wellbeing estimated that nearly half of Australians aged 16-85 years had suffered from a mental disorder during their lifetime [2]. This evidently impacts quality of life, as mental and substance use disorders were the fourth highest disease group contributing to Australia’s total burden of disease in 2015 [3].

Australians living in rural areas are reported to have a similar prevalence of mental illness compared to their urban counterparts [4]. However, they face issues exacerbated by their location, such as access to mental health services. The rate of Medicare-subsidised mental health specific services accessed by patients in 2018-19 was lower in both regional and remote areas of Australia when compared to major cities [5]. Limited access to specialised mental health care may contribute to the low usage rate. Per capita, the numbers of full-time equivalent psychiatrists, mental health nurses and psychologists in 2018 in rural areas were all lower than those of major cities [6].

Psychological distress is a widely used indicator of the mental health and well-being in population studies. The aforementioned 2017-2018 National Health Survey reported 13% of Australians over the age of 18 years experienced high or very high levels of psychological distress, an increase from 11.7% in 2014-15 [1]. However, there is limited data on psychological distress and determinants of mental health in rural communities. A 2007 study of three rural communities in South Australia and Victoria reported that 10% of respondents experienced high or very high levels of psychological distress [7]. Baseline data from a cohort study conducted in rural New South Wales (NSW) found relatively similar values of high levels of distress in respondents from inner regional (9.2%), outer regional (7.5%), remote (5.4%) and very remote areas (10.2%) [8].

There is growing evidence that social isolation is associated with poorer mental and physical health [9,10]. Social isolation has been linked with not only worse mental health outcomes, but also cardiovascular disease and increased all-cause mortality [10]. Social isolation can be measured in two ways: at the individual level, such as the social networks and activities an individual engages in, or, at the neighbourhood level where broader circumstances may impact the social connectedness of a community. An example of the latter is social fragmentation defined as the “conditions that fragment the social relationships within a neighbourhood, thereby inhibiting the levels of social cohesion available to residents” [11]. Examples of factors that increase fragmentation include a highly mobile residential population and high levels of non-family households, that lead to fewer and lower quality residential interactions. Aspects of social capital are harder to maintain in socially fragmented settings, such as trust, social norms and reciprocity [12]. Consequently, a highly fragmented neighbourhood could leave people isolated and more susceptible to mental health issues.

Recent research has shown that greater levels of social fragmentation are associated with poorer mental health [11]. There have also been several international studies undertaken to further explore this relationship. A study in England found an association between psychological distress and area-level social fragmentation in young adults [12]. An association between depressive symptoms and social fragmentation has also been observed through research in both Japan and New Zealand [13,14]. More recently, an English study found that people in more socially fragmented areas were more likely to be diagnosed with severe mental illness [15].

To measure neighbourhood social fragmentation in an Australian setting, the Australian Neighbourhood Social Fragmentation Index (ANSFI) has recently been developed [16]. Using this index, spatial variation of social fragmentation and depression was found in west Adelaide. There was also an association between the family component of social fragmentation and depression, as well as convergence in hotspots of the two variables.

Studies to date have predominantly examined the relationship between neighbourhood social fragmentation and mental health in urban populations. Further research is necessary in order to determine whether similar relationships exist in rural populations. While there are studies conducted in rural areas that have investigated social fragmentation, these have been in relation to specific mental health outcomes. A study in rural villages of China found an association with certain measures of social fragmentation and rates of suicide [17]. Additionally, a study in Scotland found a strong association with measures of social fragmentation and first-ever admission rates for psychosis, independent of urban or rural area [18]. However, knowledge is lacking about the effects of social fragmentation on a general indicator of rural mental health such as psychological distress.

The aims of this current study were to:

1. Describe the prevalence of psychological distress in the rural south-eastern NSW population.
2. Analyse the association between psychological distress and social isolation.
3. Identify any hotspots of psychological distress and social fragmentation and whether there is any convergence between the two.

## Methods

### Recruitment, participants and data collection

This study consists of a volunteer sample of adults aged 18 years or over living in rural areas within three primary healthcare networks (PHN) that include a representation of regional and remote areas: South Eastern NSW PHN, Murrumbidgee PHN and Western NSW PHN. The inclusion criterion of rurality was validated according to the remoteness area classification in the Australian Statistical Geography Standard (ASGS) [19] as those resident outside of major cities.

This study was part of a broader pilot survey on the social and cultural determinants of health in rural NSW which ran from October 2019 to April 2020. The cross-sectional online survey comprised of 56 questions on mental health, diet, cultural capital and social characteristics. The current study focused on the psychological distress component of the data collected.

Promotion of the survey was facilitated using sponsored Facebook posts and flyers placed in GP clinics affiliated with the Australian National University (ANU) Rural Clinical School, and at ANU Rural Medical Society blood pressure tents at a range of rural shows in NSW over the spring and summer.

### Psychological distress

The psychological distress assessment involved the 10-item Kessler Psychological Distress Scale, a validated measure used frequently in survey research [20]. It is included in the Australia-wide National Health Survey as a measure of psychological distress, in addition to its previous use in other Australian studies [1,7,8,21]. Items relate to the level of anxiety and depressive symptoms in the past four weeks. A response on a five-point scale is selected for each item. The K10 score is the sum of these responses, ranging from 10 (lowest psychological distress) to 50 (highest psychological distress). When deriving the K10 score, items 3 and 6 were recoded to “none of the time” if the response to the previous question was also “none of the time”, as per the standard interpretation [20]. Using previous Australian thresholds for likelihood of a clinical case, K10 scores were categorised into low (10-15), moderate (16-25) and high (26-50) groups [21].

### Social fragmentation and isolation

The ANSFI was used to measure neighbourhood-level social fragmentation [16]. The index is measured at the Statistical Area level 1 (SA1) of the Australian Statistical Geography Standard (ASGS) [19] in which the average population is approximately 400 persons. Through a principal components analysis, candidate variables were identified from the Australian Bureau of Statistics to create the ANSFI, which comprises of two distinct components labelled family (ANSFI_fam_) and mobility (ANSFI_mob_) [16]. The variables that contribute to the family component are: lone person household, non-family household with renting, and married people. The variables that contribute to the mobility component are: people living <1 year in the neighbourhood, families with school children, and people lived more than 5 years in the neighbourhood. Across the various SA1 regions of Australia, ANSFI_fam_ ranged from −4.4 to 11.6, while ANSFI_mob_ ranged from −2.8 to 15.4 [16]. Higher values indicate greater neighbourhood-level social fragmentation. In the current study, each participant address was geocoded at the SA1 level to link to ANSFI_fam_ and ANSFI_mob_ scores.

To measure social isolation at the individual level, the number of a range of occupations among the participants’ social networks was used. This measure has previously been used to record social capital [22,23]. Participants were presented with a list of 25 different occupations. The sum of the occupations in which they knew someone socially gave a score out of 25. A lower score suggests a smaller social network and an increased degree of individual-level social isolation.

### Demographic variables

Participants were asked to report a set of demographic factors including age, gender, education, marital status, employment status and annual household income. For the purpose of analysis, certain demographic variables were recoded from the original responses. Employment status was re-categorised into full-time employment, part-time employment and economically inactive. Marital status was grouped into three categories: never married, divorced/separated/widowed and married/domestic partnership.

Residential addresses were geocoded to SA1 using the 2011 ASGS in order to derive the area measures: ANSFI scores, rurality as measured by the 2011 ASGS and socioeconomic index for area using the index of relative socioeconomic advantage and disadvantage (IRSAD) developed by the Australian Bureau of Statistics [24,25]. Rurality was classified using 2011 ASGS Remoteness Areas (RA) as major cities (RA1), inner regional (RA2), outer regional (RA3), remote (RA4) and very remote (RA5) [24]. Those residing in major cities were excluded from analysis. The range of IRSAD across the whole of Australia is between 664 and 1126 [25]. Higher scores indicate less socioeconomic disadvantage.

### Relationships between social fragmentation, isolation and psychological distress

The descriptive statistics used for numerical variables were the mean, median, standard deviation and the range. For categorical variables, counts and percentages were calculated. Pearson’s correlation coefficient and scatterplots were used for initial exploration of the relationships between the K10 score and social fragmentation and isolation variables. Odds ratios (OR) and 95% confidence intervals (CI) for associations with binary indicators of psychological distress were obtained using multilevel logistic regression models to account for geographic clustering.

Data were analysed using SPSS version 24.

### Spatial analysis

ArcGIS version 10.5 was used to conduct spatial analyses and mapping. Cases were aggregated at the SA1 level to calculate psychological distress prevalence and examine spatial variation of psychological distress across the study area. However, there were not enough cases in each SA1, thus cases were subsequently aggregated at a postal area level instead [26].

Using the Global Moran’s I technique [27], the spatial autocorrelation of psychological distress and social fragmentation (both family and mobility components) was analysed. The Global Moran’s I technique is used to assess whether pattern of data is spatially clustered, dispersed or random.

### Supplementary analysis

A supplementary analysis was also performed for the occupations known socially variable, excluding the relatively large number of participants who only selected one occupation. To compare social isolation results against those for socio-economic disadvantage, the relationship between IRSAD and high psychological distress was also analysed.

### Ethics approval

The study obtained ethics approval from the Australian National University Human Ethics Committee (protocol 2019/394).

## Results

### Response, demographic and socioeconomic characteristics

There were 456 surveys submitted, of which complete data on the K10 was available for 415. Six respondents lived in major cites (RA1), while one rural respondent had missing data for the ANSFI. This left 408 respondents who had complete information for both social fragmentation and the K10, as well as resided in a rural area. Of these 408 respondents, there were 403 who also had information about occupations known socially.

The sample was predominantly female, comprising of 93% of the respondents (Table 1). Additionally, 88% of respondents were aged 40 years or over. Education level was evenly distributed, with similar numbers across postgraduate, bachelor and high school or equivalent categories. Only 15 participants (4%) did not finish high school. The majority of the sample were married or in a domestic partnership (67%). There were 40% of participants who were economically inactive and 63% of these were retired. Nearly all respondents either lived in inner regional (45%) or outer regional (53%) areas, with only seven from remote areas (2%).

**Table 1.** Sociodemographic characteristics by K10 category (n=408)

| | Total | Low (0-15) | Moderate (16-24) | High (>=25) | $\chi^2$ | p |
| --- | --- | --- | --- | --- | --- | --- |
|  | n (%) | n (%) | n (%) | n (%) |  |  |
| **Age** |  |  |  |  | 18.31 | 0.05 |
| 18-29 | 16 (3.9) | 2 (12.5) | 7 (43.8) | 7 (43.8) |  |  |
| 30-39 | 30 (7.4) | 8 (26.7) | 13 (43.3) | 9 (30.0) |  |  |
| 40-49 | 78 (19.1) | 21 (26.9) | 26 (33.3) | 31 (39.7) |  |  |
| 50-59 | 109 (26.7) | 31 (28.4) | 41 (37.6) | 37 (33.9) |  |  |
| 60-69 | 125 (30.6) | 39 (31.2) | 58 (46.4) | 28 (22.4) |  |  |
| >=70 | 47 (11.5) | 14 (29.8) | 27 (57.4) | 6 (12.8) |  |  |
| Missing | 3 (0.7) | 0 (0.0) | 2 (66.7) | 1 (33.3) |  |  |
| **Gender** |  |  |  |  | 4.54* | 0.25 |
| Male | 25 (6.1) | 9 (36.0) | 7 (28.0) | 9 (36.0) |  |  |
| Female | 380 (93.1) | 105 (27.6) | 167 (43.9) | 108 (28.4) |  |  |
| Other | 2 (0.5) | 1 (50.0) | 0 (0.0) | 1 (50.0) |  |  |
| Missing | 1 (0.2) | 0 (0.0) | 0 (0.0) | 1 (100.0) |  |  |
| **Education** |  |  |  |  | 13.61 | 0.03 |
| Postgraduate | 139 (34.1) | 45 (32.4) | 54 (38.8) | 40 (28.8) |  |  |
| Bachelor's | 143 (35.0) | 45 (31.5) | 66 (46.2) | 32 (22.4) |  |  |
| High school or equivalent | 110 (27.0) | 22 (20.0) | 44 (40.0) | 44 (40.0) |  |  |
| Did not finish high school | 15 (3.7) | 3 (20.0) | 9 (60.0) | 3 (20.0) |  |  |
| Missing | 1 (0.2) | 0 (0.0) | 1 (100.0) | 0 (0.0) |  |  |
| **Marital status** |  |  |  |  | 3.45 | 0.49 |
| Never married | 40 (9.8) | 12 (30.0) | 13 (32.5) | 15 (37.5) |  |  |
| Divorced/<br>separated/widowed | 92 (22.5) | 30 (32.6) | 37 (40.2) | 25 (27.2) |  |  |
| Married/domestic<br>partnership | 274 (67.2) | 73 (26.6) | 123 (44.9) | 78 (28.5) |  |  |
| Missing | 2 (0.5) | 0 (0.0) | 1 (50.0) | 1 (50.0) |  |  |
| **Employment status** |  |  |  |  | 2.32 | 0.68 |
| Full-time employment | 156 (38.2) | 50 (32.1) | 62 (39.7) | 44 (28.2) |  |  |
| Part-time employment | 89 (21.8) | 25 (28.1) | 39 (43.8) | 25 (28.1) |  |  |
| Economically inactive | 163 (40.0) | 40 (24.5) | 73 (44.8) | 50 (30.7) |  |  |
| **Rurality** |  |  |  |  | 1.66* | 0.80 |
| Inner regional (RA2) | 185 (45.3) | 52 (28.1) | 84 (45.4) | 49 (26.5) |  |  |
| Outer regional (RA3) | 216 (52.9) | 61 (28.2) | 87 (40.3) | 68 (31.5) |  |  |
| Remote (RA4) | 7 (1.7) | 2 (28.6) | 3 (42.9) | 2 (28.6) |  |  |
| **Annual household income** |  |  |  |  | 17.64 | 0.13 |
| Less than \$20,000 | 38 (9.3) | 6 (15.8) | 15 (39.5) | 17 (44.7) | | |
| \$20,001-\$40,000 | 60 (14.7) | 16 (26.7) | 21 (35.0) | 23 (38.3) | | |
| \$40,001-\$60,000 | 78 (19.1) | 18 (23.1) | 36 (46.2) | 24 (30.8) | | |
| \$60,001-\$80,000 | 33 (8.1) | 11 (33.3) | 14 (42.4) | 8 (24.2) | | |
| \$80,001-\$100,000 | 56 (13.7) | 19 (33.9) | 20 (35.7) | 17 (30.4) | | |
| \$100,001-\$150,000 | 81 (19.9) | 27 (33.3) | 35 (43.2) | 19 (23.5) | | |
| More than \$150,000 | 53 (13.0) | 17 (32.1) | 28 (52.8) | 8 (15.1) | | |
| Missing | 9 (2.2) | 1 (11.1) | 5 (55.6) | 3 (33.3) |  |  |

\*Fisher's exact test used as expected count <5

### Prevalence of psychological distress

The breakdown of psychological distress in the sample across the three K10 groups consisted of 115 participants (28%) in the low distress group, 174 (43%) in the moderate distress group and 119 (29%) in the high distress group.

The prevalence of psychological distress decreased with age (Table 1), although there were only a few respondents in the younger age groups and those aged 70 years or more. The prevalence of psychological distress in those that have completed a bachelor’s degree (69%) or postgraduate degree (68%) was lower compared to those with only high school or equivalent education (80%) or those that did not finish high school (80%). High psychological distress was also most common in those who were never married, lived in outer regional areas, and those on lower incomes, although the differences were not statistically significant.

### Social fragmentation, isolation and psychological distress

In the sample, ANSFI_fam_ ranged from −1.84 to 7.82, with a mean of −0.001 (median −0.13). ANSFI_mob_ ranged from −2.39 to 11.05, with a mean of −0.16 (median −0.17). The mean number of occupations known socially was 11.27 (median 11.0) and ranged from 1 to 25.

There was little evidence of a relationship found between social fragmentation or social isolation and psychological distress when analysing the K10 score as a continuous variable. The correlation coefficients were −0.01 (p=0.81) and 0.08 (p=0.11) for ANSFI_fam_ and ANSFI_mob_ respectively. When excluding three ANSFI outliers, the correlation coefficient for ANSFI_fam_ was −0.01 (p=0.77), and for ANSFI_mob_ it was 0.03 (p=0.53). The correlation coefficient between number of occupations known socially and K10 was −0.08 (p=0.13). Due to the large number of participants who only chose one occupation (n=34), the analyses were repeated excluding these participants to check for any association. However, the results were consistent with those from the full sample (data not shown).

In multilevel logistic regression models there was little evidence of associations between social fragmentation or isolation and either high or moderate/high levels of psychological distress (Table 2). The strongest association was for ANSFI_mob_, with the odds of high psychological distress increasing by 4% (95% CI 0.99-1.09) for each unit increase in the score. No association was found between IRSAD and high psychological distress or moderate to high psychological distress. However, correlation with the continuous K10 variable revealed a small inverse relationship (r=-0.12, p=0.01).

**Table 2.** Crude associations between levels of social fragmentation, isolation, socioeconomic position and psychological distress

| | High distress (K10 ≥25) | | | | | | |
| --- | --- | --- | --- | --- | --- | --- | --- |
|  | Cases (n=119) |  | Non-cases (n=289) |  | OR | 95% CI | p-value |
|  | Mean | SD | Mean | SD |  |  |  |
| ANSFI<sub>fam</sub> | -0.07 | 0.80 | 0.03 | 1.02 | 0.98 | 0.93, 1.03 | 0.51 |
| ANSFI<sub>mob</sub> | -0.05 | 1.30 | -0.20 | 0.80 | 1.04 | 0.99, 1.09 | 0.13 |
| IRSAD | 952.8 | 121.6 | 968.1 | 75.2 | 1.00 | 1.00, 1.00 | 0.14 |
| Number of occupations known socially | 10.76 | 6.73 | 11.48 | 6.43 | 1.00 | 0.99, 1.00 | 0.27 |
| | Moderate to high distress (K10 ≥16) | | | | | | |
|  | Cases (n=293) |  | Non-cases (n=115) |  | OR | 95% CI | p-value |
|  | Mean | SD | Mean | SD |  |  |  |
| ANSFI<sub>fam</sub> | 0.01 | 1.01 | -0.03 | 0.81 | 1.01 | 0.96, 1.06 | 0.72 |
| ANSFI<sub>mob</sub> | -0.13 | 1.04 | -0.21 | 0.78 | 1.02 | 0.97, 1.06 | 0.50 |
| IRSAD | 959.8 | 99.1 | 973.5 | 67.0 | 1.00 | 1.00, 1.00 | 0.15 |
| Number of occupations known socially | 10.92 | 6.57 | 12.15 | 6.34 | 0.99 | 0.99, 1.00 | 0.11 |

SD: standard deviation, OR: odds ratio, 95% CI: 95% confidence interval, ANSFI: Australian Neighbourhood Social Fragmentation Index, IRSAD: Index of Relative Socioeconomic Advantage and Disadvantage

### Spatial analysis

Figure 1 demonstrates the spatial distribution of psychological distress and the two components of social fragmentation among the postal areas of the sample. Darker shades indicate higher psychological distress or social fragmentation. For psychological distress, the Global Moran’s I indicated a random pattern of spatial distribution (−0.04, p=0.39) (Figure 2). The results were similar for social fragmentation. The Global Moran’s I indices for ANSFI_fam_ and ANSFI_mob_ were 0.001 (p=0.51) and 0.01 (p=0.49) respectively. These results show that there is no significant spatial autocorrelation of psychological distress or social fragmentation at the postal area level in the sample.

**Fig. 1.**
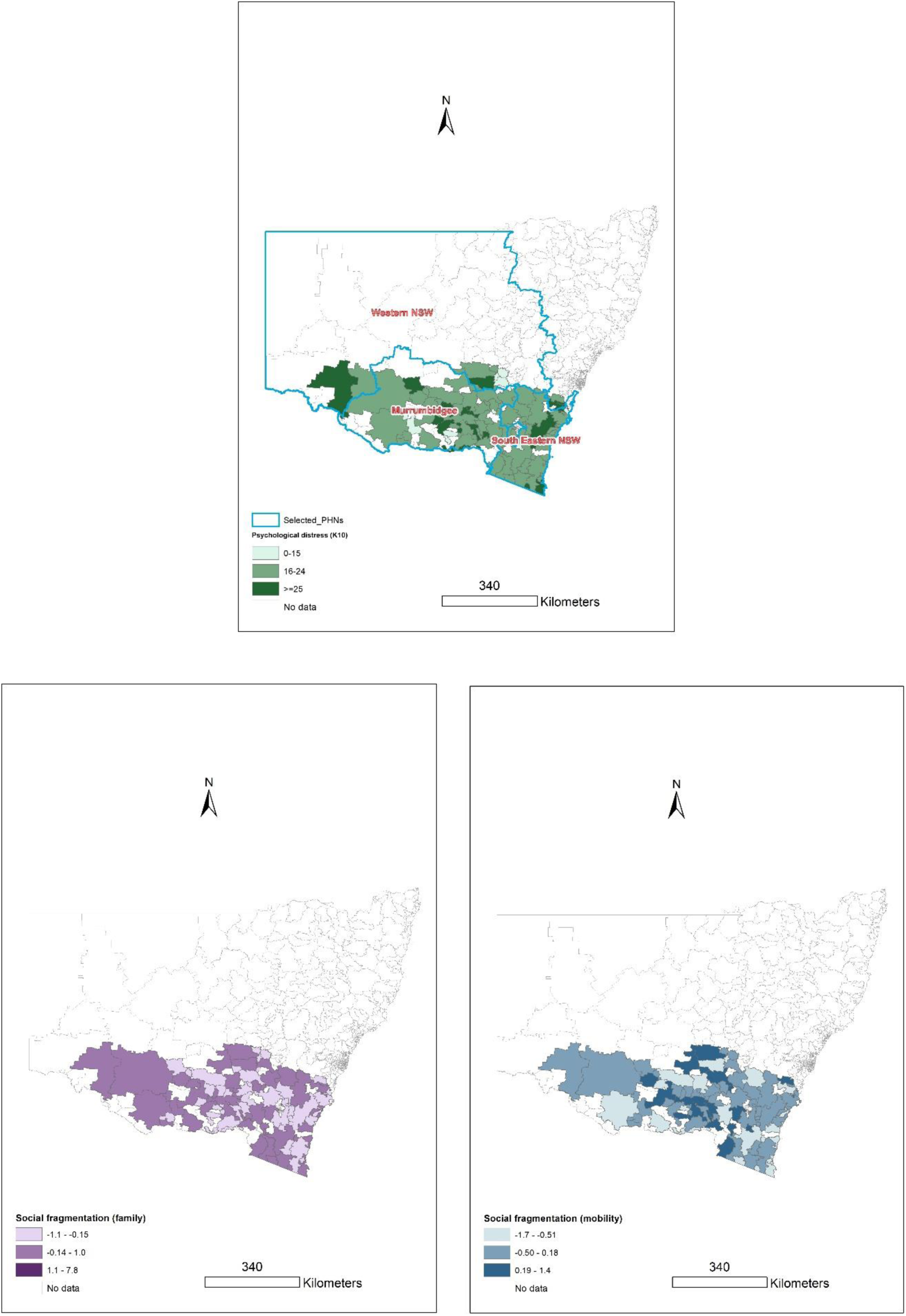
Spatial variation of psychological distress and social fragmentation components

**Fig. 2.**
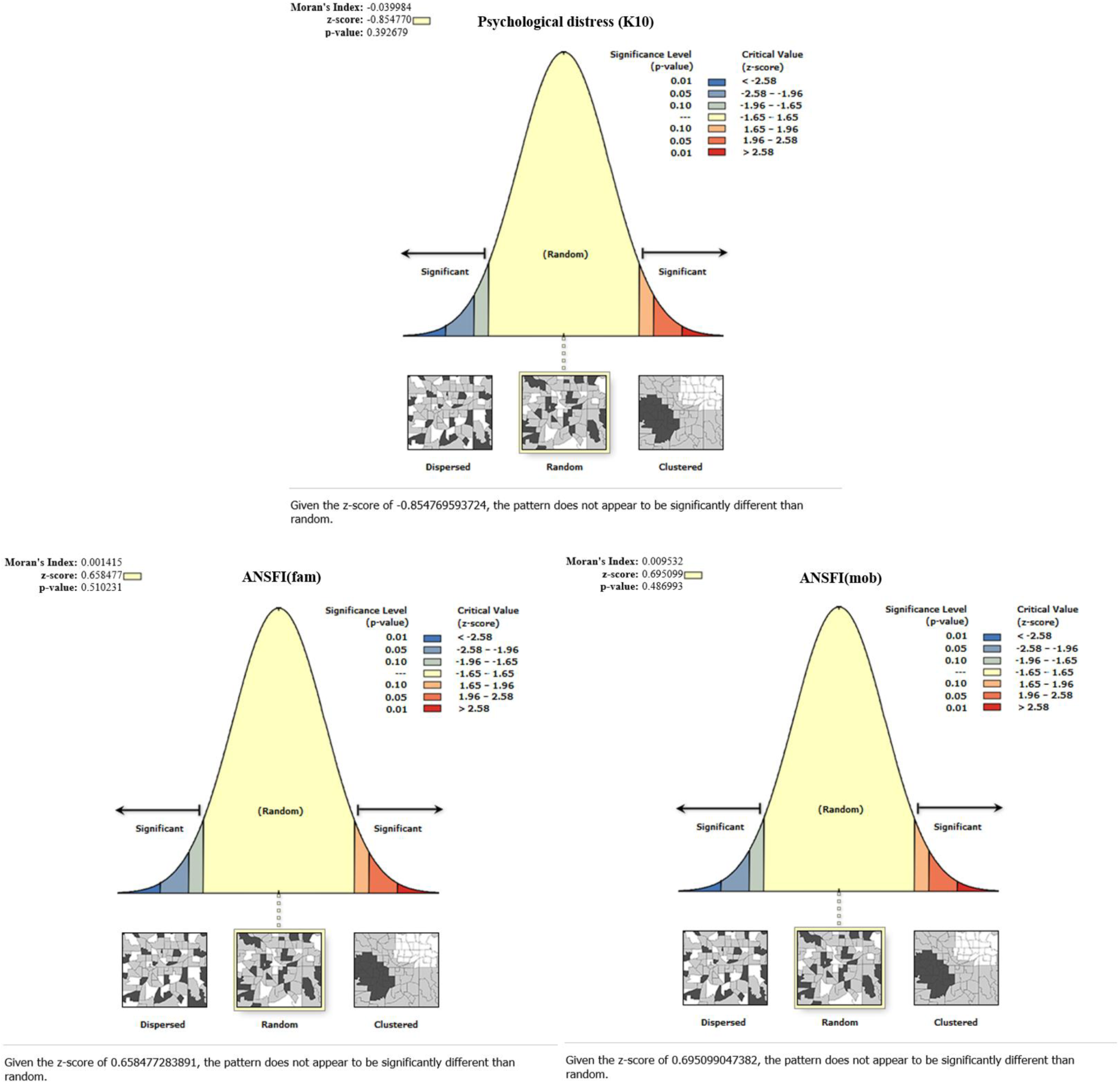
Spatial autocorrelation reports for psychological distress and social fragmentation components

## Discussion

The aim of this study was to describe the prevalence of psychological distress in south-east rural NSW and to investigate relationships with area-level social fragmentation and individual-level social isolation. While there was a relatively high level of psychological distress in the sample, there was little evidence to support an association between psychological distress and social fragmentation or isolation. Spatial analysis revealed the spread of psychological distress and social fragmentation in the sample to be randomly distributed at the postal area level with no significant clustering.

The prevalence of high psychological distress was high compared to previous studies, with 29% of the of the sample having a K10 score of 25 or higher. Only 13% of adults across the whole of Australia in the 2017 National Health Survey experienced high levels of psychological distress, [1] while an earlier study of a relatively large rural NSW cohort also had lower prevalence levels of 9.2% in inner regional areas and 7.5% in outer regional areas [8]. Both these studies also used the K10 as the outcome measure. There may be several factors contributing to the considerably high levels of psychological distress in the current sample. First, the sample was predominantly female (93%). Previous research has consistently shown a higher prevalence of depression and anxiety in females than males [28-30]. Australian data supports this, with 14.5% of women compared to 11.3% of men reporting high or very high levels of psychological distress [1]. Additionally, the timing of the survey coincided with a devastating bushfire season, as well as the start of the COVID-19 outbreak in Australia. NSW was one of the worst affected Australian states by the 2019-20 bushfires and it is likely that a considerable number of participants in the study would have been directly or indirectly affected. A survey of approximately 3000 Australians conducted in January 2020 regarding the 2019-20 bushfire season found that more than half of respondents felt anxious or worried due to the bushfires [31]. The COVID-19 pandemic has also had a profound impact on the mental health of Australians, with 78% of participants of a cross-sectional study conducted in March-April 2020 reporting that their mental health had worsened since the outbreak [32]. Lastly, the sample may be subject to selection bias, as those with higher levels of psychological distress may be more likely to respond and complete the survey.

The current research suggests that social fragmentation may not have a significant effect on psychological distress in a rural setting. Previous research using the ANSFI examined an urban area of west Adelaide [16] and our study is the first time that the index has been used in a rural population. The current results contrast with the findings in urban west Adelaide, as well as international research showing social fragmentation associations with depressive symptoms and suicide in rural areas [13,17]. Research also conducted in rural NSW suggests that individual-level attributes and perceptions are more influential than area level characteristics in relation to wellbeing in rural and remote communities [33], which may account for the lack of an observed association between social fragmentation and psychological distress. However, in this current study there was also no clear association found between psychological distress and the individual-level social isolation measure, occupations known socially.

Despite the results of the study, social isolation should still be considered in relation to rural mental health. Along with the international research regarding social fragmentation, other Australian studies have underlined the importance of social connection in rural mental health. Higher levels of social capital have been shown to be significantly associated with better mental health in South Australian rural residents [34]. In the same study, higher levels of certain social capital variables such as networks (measured as frequency of socialising and participation in community group activities) and participation in civic activities were reported in rural areas when compared with urban areas. It is possible that the measures used in the current study (neighbourhood social fragmentation and occupations known socially) might not accurately reflect levels of social isolation in rural areas. For example, while occupations known socially may represent the size of an individuals’ social network, it does not provide information about the quality of the relationships or the frequency of socialising. Further investigation could explore the effects of these other aspects of social isolation on mental health. Another hypothesis might be that higher levels of community and civic participation in rural areas may protect against any possible harmful effects of social fragmentation on mental health. Lastly, while social support may be important for rural wellbeing, other drivers of psychological distress could also play a major role. These include recent adverse events, individual dispositional factors and availability of service and support [33].

There was no significant spatial clustering for psychological distress or the components of social fragmentation. This differs from previous research using the ANSFI, where clusters of psychological distress and social fragmentation were found in urban areas of west Adelaide [16]. It is possible that clustering of these variables is less of a phenomenon in rural areas, or that there may be a more random spatial distribution at the postal area level used compared to the neighbourhood level (SA1).

There are several limitations to the study. The sample consisted of volunteers and thus was not representative of the south-eastern rural NSW population. There was a very high proportion of females in the sample (93.1%) compared to the proportions that reside in inner regional (50.9%) and outer regional (49.9%) NSW areas [35,36]. There was also an overrepresentation of participants in the 50-59 and 60-69-year-old age groups, many of whom were retired [35,36]. Additionally, the study used self-reported information as the psychological distress outcome and the individual social isolation measure. Nevertheless, while the K10 is self-reported, it is a well validated instrument and has also been used in the National Health Survey and previous Australian research [1,7,8,21]. The measure of occupations known socially may not accurately capture the complex nature of individual-level social isolation, while there were also a large number of participants who only chose one occupation. This may be due to misinterpretation of the instructions, or it is also possible that participants truly only knew one person from the occupations listed. Finally, due to the sample size, spatial analysis could only take place at the postal area level, rather than the SA1 area as originally planned. Further analysis at a rural small area level may provide a greater insight into possible clustering and hotspots of social isolation and mental illness.

The results from this study suggest that social fragmentation or isolation may not have a significant effect on psychological distress in a rural setting. Further investigation of the effects of social isolation and social fragmentation in rural areas is warranted. A deeper knowledge of how social isolation affects mental health in rural communities at both an individual and area-level will facilitate more effective and targeted interventions to protect and improve mental health in different settings.

## Data Availability

Data will be archived following publication of outputs through the ANU Data Repository. Access to data requires permission of the data custodian.

## Acknowledgements

We thank Dr. Xiaozhou Zhang for assistance with co-development of the questionnaire. We acknowledge the help of ANU Medical School, ANU Rural Clinical School, and ANU Rural Medical Society for promoting the survey on Facebook and Ms. Evangeline Wong for promoting the survey at Rural Shows and on local news radio.

## Declarations

### Consent to participate

All persons gave their informed consent prior to their inclusion in the study.

### Author contributions

All authors contributed to the study conception and design. Material preparation, data collection and analysis were performed by Victor Forcadela, Claudia Slimings and Nasser Bagheri. The first draft of the manuscript was written by Victor Forcadela and all authors commented on previous versions of the manuscript. All authors read and approved the final manuscript.

## References

1. Australian Bureau of Statistics (2018) National Health Survey: First results, 2017-18. https://www.abs.gov.au/statistics/health/health-conditions-and-risks/national-health-survey-first-results/2017-18. Accessed 26 September 2020

2. Australian Bureau of Statistics (2008) National Survey of Mental Health and Wellbeing: Summary of Results, 2007. https://www.abs.gov.au/statistics/health/mental-health/national-survey-mental-health-and-wellbeing-summary-results/latest-release. Accessed 30 September 2020

3. Australian Institute of Health and Welfare (2020) Burden of disease. https://www.aihw.gov.au/reports/australias-health/burden-of-disease. Accessed 11 August 2020

4. Australian Institute of Health and Welfare (2019) Rural & remote health, Health status and outcomes. https://www.aihw.gov.au/reports/rural-remote-australians/rural-remote-health/contents/health-status-and-outcomes. Accessed 12 August 2020

5. Australian Institute of Health and Welfare (2020) Mental health services in Australia, Mental health-specific services. https://www.aihw.gov.au/reports/mental-health-services/mental-health-services-in-australia/report-contents/medicare-subsidised-mental-health-specific-services/mental-health-specific-services. Accessed 12 August 2020

6. Australian Institute of Health and Welfare (2020) Mental health services in Australia, Mental health workforce. https://www.aihw.gov.au/reports/mental-health-services/mental-health-services-in-australia/report-contents/mental-health-workforce. Accessed 12 August 2020

7. Kilkkinen A, Kao-Philpot A, O’Neil A, Philpot B, Reddy P, Bunker S, Dunbar J (2007) Prevalence of psychological distress, anxiety and depression in rural communities in Australia. Aust J Rural Health 15 (2):114–119. doi:10.1111/j.1440-1584.2007.00863.x

8. Kelly BJ, Stain HJ, Coleman C, Perkins D, Fragar L, Fuller J, Lewin TJ, Lyle D, Carr VJ, Wilson JM, Beard JR (2010) Mental health and well-being within rural communities: the Australian rural mental health study. Aust J Rural Health 18 (1):16–24. doi:10.1111/j.1440-1584.2009.01118.x

9. Holt-Lunstad J (2018) Why Social Relationships Are Important for Physical Health: A Systems Approach to Understanding and Modifying Risk and Protection. Annu Rev Psychol 69:437–458. doi:10.1146/annurev-psych-122216-011902

10. Leigh-Hunt N, Bagguley D, Bash K, Turner V, Turnbull S, Valtorta N, Caan W (2017) An overview of systematic reviews on the public health consequences of social isolation and loneliness. Public Health 152:157–171. doi:10.1016/j.puhe.2017.07.035

11. Ivory VC, Collings SC, Blakely T, Dew K (2011) When does neighbourhood matter? Multilevel relationships between neighbourhood social fragmentation and mental health. Soc Sci Med 72 (12):1993–2002. doi:10.1016/j.socscimed.2011.04.015

12. Fagg J, Curtis S, Stansfeld SA, Cattell V, Tupuola AM, Arephin M (2008) Area social fragmentation, social support for individuals and psychosocial health in young adults: evidence from a national survey in England. Soc Sci Med 66 (2):242–254. doi:10.1016/j.socscimed.2007.07.032

13. Takagi D, Kondo K, Kondo N, Cable N, Ikeda K, Kawachi I (2013) Social disorganization/social fragmentation and risk of depression among older people in Japan: multilevel investigation of indices of social distance. Soc Sci Med 83:81–89. doi:10.1016/j.socscimed.2013.01.001

14. Pearson AL, Ivory V, Breetzke G, Lovasi GS (2014) Are feelings of peace or depression the drivers of the relationship between neighbourhood social fragmentation and mental health in Aotearoa/New Zealand? Health Place 26:1–6. doi:10.1016/j.healthplace.2013.11.002

15. Grigoroglou C, Munford L, Webb RT, Kapur N, Ashcroft DM, Kontopantelis E (2020) Prevalence of mental illness in primary care and its association with deprivation and social fragmentation at the small-area level in England. Psychol Med 50 (2):293–302. doi:10.1017/s0033291719000023

16. Bagheri N, Batterham PJ, Salvador-Carulla L, Chen Y, Page A, Calear AL, Congdon P (2019) Development of the Australian neighborhood social fragmentation index and its association with spatial variation in depression across communities. Soc Psychiatry Psychiatr Epidemiol 54 (10):1189–1198. doi:10.1007/s00127-019-01712-y

17. Li LW, Xu H, Zhang Z, Liu J (2016) An Ecological Study of Social Fragmentation, Socioeconomic Deprivation, and Suicide in Rural China: 2008-2010. SSM Popul Health 2:365–372. doi:10.1016/j.ssmph.2016.05.007

18. Allardyce J, Gilmour H, Atkinson J, Rapson T, Bishop J, McCreadie RG (2005) Social fragmentation, deprivation and urbanicity: relation to first-admission rates for psychoses. Br J Psychiatry 187:401–406. doi:10.1192/bjp.187.5.401

19. Australian Bureau of Statistics (2020) Australian Statistical Geography Standard (ASGS). https://www.abs.gov.au/websitedbs/D3310114.nsf/home/Australian+Statistical+Geography+Standa rd+(ASGS). Accessed 8 August 2020

20. Andrews G, Slade T (2001) Interpreting scores on the Kessler Psychological Distress Scale (K10). Aust N Z J Public Health 25 (6):494–497. doi:10.1111/j.1467-842x.2001.tb00310.x

21. Fragar L, Stain HJ, Perkins D, Kelly B, Fuller J, Coleman C, Lewin TJ, Wilson JM (2010) Distress among rural residents: does employment and occupation make a difference? Aust J Rural Health 18 (1):25–31. doi:10.1111/j.1440-1584.2009.01119.x

22. Burgis-Kasthala S, Slimings C, Smith M, Elmitt N, Moore M (2019) Social and community networks influence dietary attitudes in regional New South Wales, Australia. Rural Remote Health 19 (3):5328. doi:10.22605/rrh5328

23. Sheppard J, Biddle N (2017) Class, capital, and identity in Australian society. Aust J Polit Sci 52 (4):500–516. doi:10.1080/10361146.2017.1364342

24. Australian Bureau of Statistics (2018) 1270.0.55.005 - Australian Statistical Geography Standard (ASGS): Volume 5 - Remoteness Structure, July 2011. https://www.abs.gov.au/AUSSTATS/abs@.nsf/DetailsPage/1270.0.55.005July%202011?OpenDocument. Accessed 8 August 2020

25. Australian Bureau of Statistics (2013) 2033.0.55.001 - Census of Population and Housing: Socio-Economic Indexes for Areas (SEIFA), Australia, 2011. https://www.abs.gov.au/ausstats/abs@.nsf/Lookup/by%20Subject/2033.0.55.001~2011~Main%20Features~Main%20Page~1. Accessed 28 August 2020

26. Australian Bureau of Statistics (2016) 1270.0.55.003 - Australian Statistical Geography Standard (ASGS): Volume 3 - Non ABS Structures, July 2016. Accessed 1 August 2020

27. Moran PA (1950) Notes on continuous stochastic phenomena. Biometrika 37 (1-2):17–23

28. Baxter AJ, Scott KM, Vos T, Whiteford HA (2013) Global prevalence of anxiety disorders: a systematic review and meta-regression. Psychol Med 43 (5):897–910. doi:10.1017/s003329171200147x

29. Culbertson FM (1997) Depression and gender. An international review. Am Psychol 52 (1):25–31. doi:10.1037//0003-066x.52.1.25

30. Kuehner C (2017) Why is depression more common among women than among men? Lancet Psychiatry 4 (2):146–158. doi:10.1016/s2215-0366(16)30263-2

31. ANU Poll 2020: Bushfires, The Environment, and Optimism For The Future (2020) ADA Dataverse. http://dx.doi.org/10.26193/S1S9I9.

32. Newby JM, O’Moore K, Tang S, Christensen H, Faasse K (2020) Acute mental health responses during the COVID-19 pandemic in Australia. PLoS One 15 (7):e0236562. doi:10.1371/journal.pone.0236562

33. Kelly BJ, Lewin TJ, Stain HJ, Coleman C, Fitzgerald M, Perkins D, Carr VJ, Fragar L, Fuller J, Lyle D, Beard JR (2011) Determinants of mental health and well-being within rural and remote communities. Soc Psychiatry Psychiatr Epidemiol 46 (12):1331–1342. doi:10.1007/s00127-010-0305-0

34. Ziersch AM, Baum F, Darmawan IG, Kavanagh AM, Bentley RJ (2009) Social capital and health in rural and urban communities in South Australia. Aust N Z J Public Health 33 (1):7–16. doi:10.1111/j.1753-6405.2009.00332.x

35. Australian Bureau of Statistics (2017) 2016 Census QuickStats: Inner Regional Australia (NSW). https://quickstats.censusdata.abs.gov.au/census_services/getproduct/census/2016/quickstat/RA11?opendocument. Accessed 15 September 2020

36. Australian Bureau of Statistics (2017) 2016 Census QuickStats: Outer Regional Australia (NSW). https://quickstats.censusdata.abs.gov.au/census_services/getproduct/census/2016/quickstat/RA12?opendocument. Accessed 15 September 2020

